# Price and Tax Elasticity of Alcohol Consumption: Evidence from Seven Indian States

**DOI:** 10.1101/2022.03.25.22272940

**Authors:** Samarth Gupta, Soumi Roy Chowdhury, Sanjib Pohit, Fikru Tesfaye Tullu, Pradeep Joshi

**Affiliations:** Amrut Mody School of Management, Ahmedabad University; The George Washington University, USA; NCAER; World Health Organization (WHO) in India

**Keywords:** Public Health, Health and Economic Development, Taxation

## Abstract

In this paper, we develop a novel methodology for constructing price indices of India Made Foreign Liquor (IMFL), which account for incompatible state-level taxation structure and policies across seven states in India. We use this price index to compute price and tax elasticity of IMFL in the seven states. We find that a 1% increase in price leads to only a 0.057% decline in consumption while a 1% increase in tax reduces alcohol consumption by 0.14%. We extend our estimates to show that a 10% increase in taxes will lead to an 8.4% increase in tax revenue. Our results are the most updated estimates of alcohol consumption elasticities, which overcomes several challenges such as varying manufacturing costs across states and varying tax across types of products within states, which other papers do not.

## 1. Introduction

The subject of alcohol holds a special significance in Indian public policy due to the dilemma policymakers face with the boons and banes associated with alcohol. Unsafe use of alcohol, on one hand, brings in significant health care burden and productivity loss (Chaloupka et al., 2019), while on the other hand, alcohol revenues serve as a major source of income generation for most Indian states (Dhanuraj & Kumar, 2014). This fiscal dependence prevents the states from imposing stronger restrictions on alcohol production and distribution.

Though overwhelming evidences suggest that alcohol consumption is highly correlated with negative health consequences (Lim et al., 2012; Whiteford et al., 2013; Allen et al., 2009; Fedirko et al., 2011), a complete ban on alcohol remains undesirable. This is due to the unintended consequences of black marketing and illicit alcohol production that a ban would give rise to. Policymakers, therefore resort to increasing alcohol prices through tax instruments so that consumption can be curbed (Elder et al., 2010) while revenue streams are kept intact.

Taxation can serve this dual purpose given the nature of the elasticity of the good, which in this case is alcohol. It is important for the policy makers to be informed of the price sensitivity of alcohol demand in India. With that objective in mind, we studied alcohol taxation structure of Indian Made Foreign Liquor (IMFL) across seven Indian states; namely Himachal Pradesh, Madhya Pradesh, Sikkim, Bihar, Karnataka, Maharashtra, and Delhi.

A multi-state analysis of alcohol taxation in India poses a variety of challenges. Alcohol items do not come under the ambit of Goods and Service Tax of the Central government. Each state has its unique alcohol taxation structure set and regulated by respective state. The principle on which the tax rates are decided vary greatly across states. For some states, the excise duties are levied on the total volume of liquor, whereas for others, the rates are based on the alcohol content. Most importantly, the lack of uniformity in data across different states makes it inherently difficult to arrive at a country wide representative price for alcohol. As Rahman (2004) rightly pointed out, all these constraints stand in the way of efficient research on price elasticity of alcohol (PED) in India.

The most comprehensive study in this subject was conducted by Mahal (2000). Using a survey of 33,000 households across 15 major states of India, Mahal found that for moderate to heavy drinkers, the elasticity is -1.00 among individuals of 15-25 years and - 0.50 for individuals aged 25 years and above. We make the following four significant departures from (Mahal, 2000) and in turn they are our contributions to the existing stream of literature. First (Mahal, 2000) computed alcohol prices under the assumptions of equal marginal cost of production of alcohol across states. This is unlikely to hold true due to the differential labour and capital costs across states. We have collected state specific Ex-Distillery Prices to accommodate for the variation in marginal cost. Second, we departed from Mahal’s assumption of one rate of taxation for different liquor categories and alternatively accounted for varying tax rates across different ranges and types of IMFL. Third, Mahal proxies the ‘average’ excise tax per unit of alcohol consumption from the ratio of total consumption with the number of state consumers of alcohol. This can be misleading as the numerator will be inflated for states with high cross-border consumption and tourism. By contrast, we compile actual tax information by visiting state excise departments. Finally, Mahal considered a binary dependent variable which was derived from frequency of consumption using 1994 data; whereas we use the level of consumption from 2011 NSSO data and control for the distribution of consumers over the price range.

## 2. Data and Measures

Given the data constraints as outlined in the previous section, gathering information on alcohol taxation required in-person visits to different state excise departments. Other than the national capital (Delhi), the six states included within the scope of the paper represent six different regions of the country—Himachal Pradesh (HP) [North]; Odisha [East]; Sikkim [North East]; Madhya Pradesh (MP) [Centre]; Karnataka [South]; and Maharashtra [West]. These states together represent 30% of the total population of India in 2011. According to a report published by the Ministry of Social Justice and Empowerment, prevalence of alcohol consumption in Delhi, Sikkim, MP lie above the national average, whereas HP, Karnataka, Bihar, and Maharashtra are below the national average.

Tax rates levied on IMFL are collated by visiting each state excise departments. Survey personnel visited excise departments of the selected states during November and December 2019 and met excise commissioners and/or other relevant officials to collect excise duty rates, margins and price data of alcohol products. For the purpose of this paper we only use the tax data of 2011. This was done to accommodate for the fact that the latest NSSO consumption rounds is available for 2011. There are no national level alcohol consumption figures available for the country after 2011. Data was shared to the survey personnel in various formats including hard copy, soft copy, and website information. Excise revenue data was collected from budget documents of the selected states.

### 2.1 Price Data

In calculating the price, three survey measures are used; (a) Ex-Warehouse Price (EWP), also known as the manufacturer’s price. EWP reflects the marginal cost of producing alcohol, (b) Excise duties (ED) and (c) Additional excise duties (AED), both of which are indirect taxes levied on alcohol and varies by states and by EWP. Before we arrived at a unique price of IMFL for a particular state, heterogeneity across states in the units of taxation are made uniform. We normalized the EWP, ED and AED across states such that the amount is represented in per litre terms.

### 2.2 Consumption Data

NSSO’s household consumer expenditure surveys provide state-level representative data on household consumption for various food and non-food items with a 30 day reference period for most items. These include litres and expenditure on alcoholic products (toddy/arrack, beer and wine separately) as well. The category of wine corresponds to IMFL for which we collected prices for the seven states. Other datasets such as NFHS and IHDS only record the frequency of alcoholic products consumed and not the actual amount. Therefore, we rely on NSSO to compute elasticity measures. For the current study, we use NSSO’s 68^th^ round conducted in 2011-12 (NSSO 2011-12 from hereon), which is the last conducted publicly available household consumer expenditure survey.

### 2.3 Descriptive Statistics

Table 1 below shows the monthly IMFL consumption of households derived from NSSO 2011-12. The consumption amount ranges from 0.93 litres in Sikkim to 1.66 litres in Himachal Pradesh. There is considerable heterogeneity within state in alcohol consumption as evident from the standard deviation of 2.26 for Himachal Pradesh but only 0.47 for Sikkim. Overall, we see that the self-reported mean consumption levels for households are just above 1 litre for most of the states.

**Table 1:**
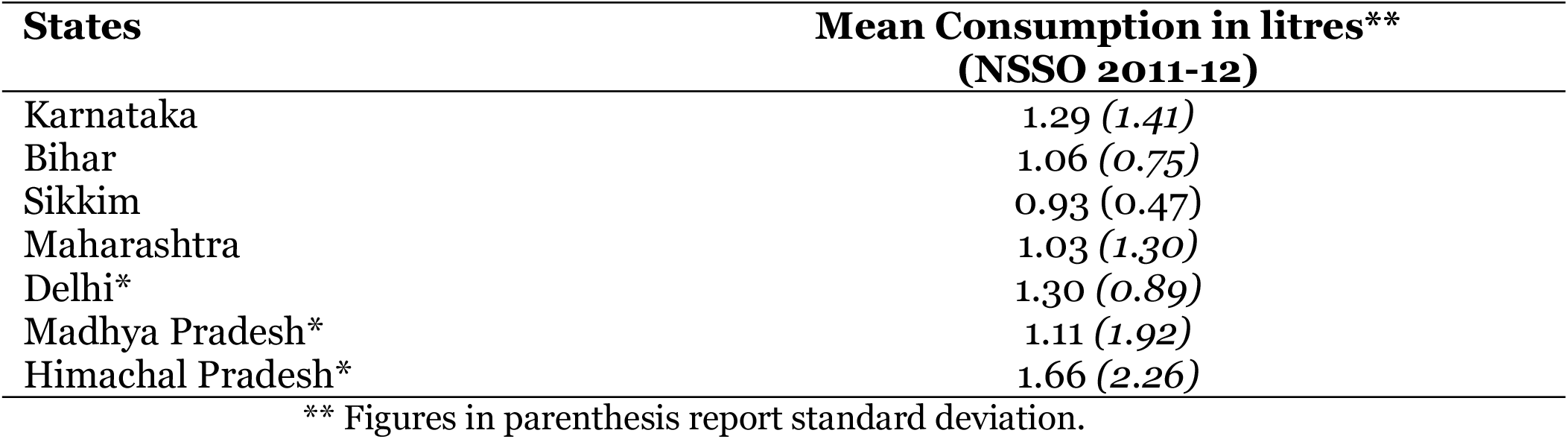
Mean consumption levels by states (NSSO 2011-12)

Delving deeper into the patterns of consumption across states, Table 2 below gives the share of monthly expenditure on alcohol over aggregate monthly expenditure for households at each quintile. We divided our households into equally-sized expenditure quintile groups of households for different states. The quintile groups are for all-India level. Then we use those intervals to see consumption expenditure for different states. The ratio of expenditure by top and bottom quintile (Q5/Q1) provides an indicator of disparity in alcohol expenditure. As we can see below, households in top quintile spend at least 2 times more than households in bottom quintile, with disparity highest in Delhi and lowest in Karnataka and Sikkim. Also, interestingly, it is Karnataka, where poorest households (at income quintile Q1) spend more than 5% of their aggregate expenditure on alcohol, as opposed to MP where this ratio is only 3.5%. For households at income quintile Q2, the percentage is even higher (7.05%). Even in absolute terms, Q1 households in Karnataka spend more than double the average of all the rest six states. This may have a bearing with the ban of arrack in Karnataka in 2006-07 which was predominantly a drink of the poorer households. The pattern of expenditure may be suggestive of the fact that people, otherwise who would consume arrack have switched to consuming a relatively expensive liquor IMFL post arrack ban.

**Table 2:** Monthly Expenditure on Alcohol as a share of Aggregate Monthly Expenditure (NSSO 211-12)

|  | Q1 | Q2 | Q3 | Q4 | Q5 |
| --- | --- | --- | --- | --- | --- |
| Himachal Pradesh | 2.92 | 3.48 | 3.80 | 3.50 | 4.31 |
| Bihar | 3.06 | 1.99 | 4.61 | 3.26 | 2.30 |
| Delhi | - | 1.98 | 4.04 | 2.16 | 3.33 |
| Sikkim | 3.09 | 4.59 | 4.21 | 4.82 | 3.81 |
| Madhya Pradesh | 3.50 | 3.08 | 3.68 | 4.05 | 4.01 |
| Maharashtra | 3.36 | 4.12 | 3.71 | 4.89 | 3.14 |
| Karnataka | 5.07 | 7.05 | 5.97 | 4.87 | 4.16 |
| All Seven States | 3.41 | 3.72 | 4.61 | 4.24 | 3.67 |

NCAER estimates using data from NSSO 2011-12
**Note:** No Delhi household in the lowest quintile group reported expenditure on alcohol.
**Source:** Authors' estimate.
Q1-Q5 are Quintile groups according to aggregate expenditure for India. Q1 is the lowest quintile (poorest households) and Q5 is the highest quintile group (richest households).

## 3. Methods

### 3.1 Price calculation

The normalized Excise duty, Additional Excise duty, Warehouse prices are used in a two-step pricing methodology to arrive at an effective prices for each of the seven states. The price calculation involves the following.

The first step is calculating the Unweighted Prices using the below Equation (1)

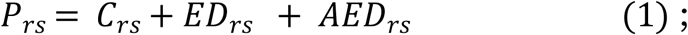

*P*_*rs*_ is the Unweighted Prices calculated for each range (r) of IMFL in state (s) ; *C*_*rs*_ is the marginal cost (EWP) of producing IMFL; *ED*_*rs*_ is the rate of excise duties levied by the states, and *AED*_*rs*_ is the additional excise duties. There are different ranges of Ex-warehouse prices for variety of IMFL in a particular state for a particular year. The ranges are to be read as cheap category of liquor versus expensive liquors. After we obtain an unweighted price for each range of IMFL in each state, in the next step, we weighted those price ranges (*P*_*rs*_) with the distribution of population consuming IMFL in that price range for that state in 2011. The information on the population proportion is obtained from NSSO 2011-12 data. We have used the dated NSSO data which is also the latest one with the underlying assumption that the shape of the distribution of alcohol consumers have not changed over the years. There might have been a shift in the distribution but the overall shape has remained constant Therefore, the Effective prices (*EP*_*S*_) of IMFL in a state for a year is a summation of all the weighted *P*_*rs*_ as shown below in Equation (2). Through this methodology we arrive at one effective price for one state for the year 2011. Also, important to note here is that for states where tax information is available for year 2011, effective prices were computed directly from the formula above. However, for states that have information for years other than 2011, we were required to inflate/ deflate the prices to represent them in 2011 prices, i.e effective prices *EP*_*S*_ were adjusted with CPI IMFL to represent them in 2011 prices. Appendix 2 provides a technical note on inflation adjustment of this price.

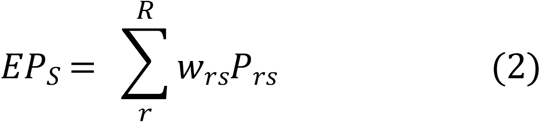

*EP*_*S*_ is the Effective (Weighted) Price for all ranges of the alcohol type in 2011

*w*_*rs*_ is the proportion of people consuming alcohol in range *P*_*rs*_ obtained from NSSO 2011.

### 3.2 Calculating Price and Tax Elasticity for seven states

The prices calculated above is then used to estimate the price and tax elasticity of IMFL for the seven states under study. In the following Equation 3, we proceed with estimating the price elasticity of alcohol consumption using a log-linear demand functional form:

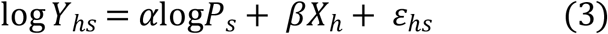

where, log*Y*_*hs*_ is regressed on log of prices in state s (log*P*_*s*_). *X*_*h*_ are the household level characteristics that may determine consumption of alcohol. These include the Education levels of household head (below primary, primary, secondary, higher secondary and above), dummy variables for religion of the household head (Hindu, Muslim, Christian, Sikh, Jain, Buddhism, and Others), social groups and ethnicity (General, SC, OBC, Others), and finally the income quintile groups of the households.

The above demand function can be derived from a household maximizing a Cobb-Douglas utility functional form subject to its income level (proxied by income quintile) and preference parameters (proxied by household characteristics such as caste, education, etc.). Elasticity is then estimated by Ordinary Least Square Estimate of *α*.

Tax elasticity is estimated by Equation 4:

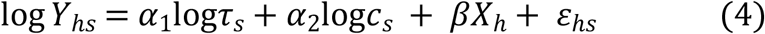

where, *τ*_*s*_ and *c*_*s*_ are tax and warehouse prices, respectively, in state s, as explained in Table 1. Using *c*_*s*_ as a covariate allows controlling for supplier conditions in the state. Then, *α*_1_ records how a 1% change in state-level tax changes alcohol consumption in the household, holding constant household characteristics (*X*_*h*_), state-specific manufacturing costs (*c*_*s*_).^3^

### 3.3 Calculating state specific price elasticity

Our price calculation led us to one price of IMFL for one state. This definitely does not mirror the actual world where areas of residence (rural versus urban), types of IMFL, price competitions, commercial or residential consumption, leads to price variation within states. Therefore to arrive at a state specific elasticity of alcohol, we resorted to the household level IMFL consumption and expenditure data obtained from NSSO 2011-12.

The unit prices vary across households in a state which is then used to estimate seven separate weighted regressions for seven states individually. Using the similar specification as in Equation 4, log of quantity consumed is regressed on the log of prices and other control variables.

## 4. Results

### 4.1 Effective Prices of IMFL

In Table 4, we present the effective prices of IMFL obtained from the two-step pricing methodology and the effective tax rates. In addition, we provide the weighted price of IMFL as reported in NSSO 2011-12 data. The NSSO price serves as a comparison and shows the relevance of our pricing methodology.

**Table 4:** Prices and taxes by states in 2011.

| States | Our Pricing Method |  | NSSO Prices<br>(Mean) |
| --- | --- | --- | --- |
|  | Effective IMFL Prices (Rs)<br>(Our Methodology) | Taxes<br>(Rs) |  |
| Karnataka | 475 | 313 | 426 |
| Bihar | 410 | 344 | 342 |
| Sikkim | 245 | 99 | 278 |
| Maharashtra* | 446 | 284 | 674 |
| Himachal Pradesh* | 256 | 167 | 319 |
| Madhya Pradesh* | 367 | 281 | 459 |
| Delhi* | 743 | 353 | 518 |

Sources: Computed by authors;
**Note:** In *Our Methodology*, the Effective Prices are calculated based on the two step methodology. We have prices for Karnataka, Bihar, Sikkim, and Maharashtra for the year 2011. For Himachal Pradesh, Madhya Pradesh, and Delhi, Prices were adjusted by CPI IMFL to 2011. NSSO prices are the mean price of IMFL calculated from the unit level NSSO consumption and expenditure data.

We see that tax rates comprise a major component of the prices. About 40%-80% of the effective price is tax alone. The marginal cost of producing alcohol consists of only a fraction of the final price. Interestingly, the NSSO prices are not very far off from what we had computed. In both the pricing methodologies, it is Sikkim where IMFL is seen to be the cheapest (Rs 245 versus Rs 278) followed by Himachal Pradesh (Rs 256 versus Rs 319). IMFL is the most expensive in Maharashtra and Delhi.

### 4.2 Price Elasticity and Tax Elasticity of seven states

Table 6 provides estimates of price elasticity from Equation 4 for different models. It is the elasticity of all the seven states taken together. Model 1 and Model 2 are two different specifications, where the former reports elasticity measure without including any control variables. Understanding that this might create omitted variable bias the latter model is run with household control variables. Price elasticity of demand for IMFL for all seven states is found to be -0.057; i.e a 10% increase in price will reduce consumption by 0.57%.

**Table 6:** Price Elasticity.

| Variables | Model 1 | Model 2 |
| --- | --- | --- |
| Price Elasticity | 0.009***<br>(0.002) | -0.057***<br>(0.002) |
| Household Controls | No | Yes |
| Observations | 2673752 | 2673752 |

\*, \*\*, \*\*\* denote significance at 10%, 5%, 1% respectively. Figures in parenthesis reports standard errors. Regressions are weighted. Controls include religion, caste, and expenditure quintile of households and size of households

Table 7 provides estimate of tax elasticity, i.e decrease in IMFL consumption for a 1% increase in taxes. Analogous to Table 4, Model 1 and 2 provide tax elasticity using the two methods with and without household controls, respectively. In model 1, tax elasticity of consumption is -0.28. In model 2, we add household characteristics which determine IMFL consumption. Now, elasticity is -0.145; a 10% increase in tax rate reduces alcohol consumption by 1.4%

**Table 7:** Tax Elasticity.

|  | Model 1 | Model 2 |
| --- | --- | --- |
| Tax Elasticity | -0.28***<br>(0.002) | -0.14***<br>(0.003) |
| Household Controls | No | Yes |
| Observations | 2673752 | 2673752 |

\*, \*\*, \*\*\* denote significance at 10%, 5%, 1% respectively. Figures in parenthesis reports standard errors. Regressions are weighted Controls include religion, caste, and expenditure quintile of households and size of households

**Table 8:** State specific Price Elasticity: With Controls.

|  | Karnataka | Bihar | Sikkim | Maharashtra | Himachal Pradesh | Madhya Pradesh | Delhi |
| --- | --- | --- | --- | --- | --- | --- | --- |
| **Price** | -0.81***<br>(0.001) | -0.10***<br>(0.002) | -0.19***<br>(0.001) | -0.24***<br>(0.002) | -0.62***<br>(0.004) | -0.35***<br>(0.002) | -0.007*<br>(0.004) |
| **N** | 1561928 | 282900 | 33,234 | 280364 | 154873 | 219599 | 140851 |

**Source:** Authors' estimate from NSSO 2011-12

Tax elasticity measure is useful to construct implications on tax revenue of an increase in tax rate. Consider the following equation for Tax Revenue:

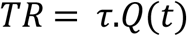

where, *τ* is tax rate and *Q* is the quantity of alcohol consumed. Consider an increase in tax by 10%. Now, the new tax is *τ*′ = 1.10 *τ*. Given tax elasticity of -0.14, quantity consumed now will be 1.4% lower.; i.e. *Q*′ = 0.986 ∗ Q(t). Now, new tax revenue will be:

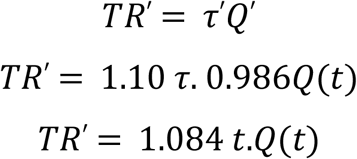

Thus, change in tax revenue can be written as:

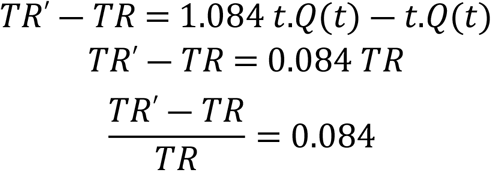

Thus, a 10% increase in tax rate increases tax revenue by 8.4%.

### 4.3 State Specific Elasticity

Finally, we compute the state specific elasticity using prices computed from expenditure and consumption form NSSO unit level data. Our overall conclusion on IMFL being an inelastic good remained the same irrespective of the methodology used. However, for the state specific measures, the values increased in magnitude. IMFL is least elastic in Delhi (−0.007) which may partly be explained by higher incomes in the state leading to high affordability. On the other hand, consumers in Karnataka (−0.81) and Himachal Pradesh (−0.62) are more price sensitive.

## 5. Conclusion

Incompatibility of alcohol taxation structures across states in India limit conducting policy-relevant exercises such as estimating price and tax elasticity of alcohol products. In this paper, we developed a novel two-step procedure to arrive at a weighted effective price of IMFL for seven states in India. Our price index accounts for manufacturing prices, ED, AED for different categories of IMFL and weights them by the distribution of households consuming IMFL at a given price range in each state. The former price metric allows us to observe the tax component separately which is useful for estimating tax elasticity. The unit prices from NSSO 2011-12 provide price variation across households within states.

Using our state-specific measure of prices, the demand for alcohol consumption is found to be highly inelastic with only a 0.057% decline in consumption due to a 1% increase in price. Similarly, an increase in tax by 1% reduces alcohol consumption by 0.14%. Tax elasticity of consumption is constructed to understand the implications of an increase in tax rate on the amount of revenue generated. We find that a 10% increase in tax rate increases revenue by 8.4%. However, it should be acknowledged that these elasticity measures reflect aggregate consumer behavior in the seven states.

We augmented our analysis by estimating state-specific price elasticity using unit prices from NSSO 2011-12 consumption and expenditure data. Richer states like Delhi have much lower elasticity perhaps owing to affordability, whereas consumers in Karnataka are more price sensitive. Understanding this variation can help on devise more impactful policy measures.

In the process of the paper, we also learnt about the dissonance on data across states. Not only the tax rates but the structure in which data is compiled also varies by states. For instance, some states report ex-warehouse prices of their product whereas others do not report any such manufacturing cost forcing the researcher to use an appropriate proxy of the manufacturing cost. Additionally, states also vary in tax regimes; i.e. some states tax alcohol by total volume while others impose a tax on alcohol content. A centralized data compilation platform ensuring data comparability should be devised so that in-depth socio-economic studies on alcohol use and related implications can be carried out.

Given a substantial amount of state revenue is generated from these taxes, it is important that the revenue is allocated to awareness campaigns and implementing other SAFER intervention as recommended by WHO^4^. Other state-specific policies including enforcement of minimum legal age of drinking, prohibition and alcohol distribution model may have a significant impact on the price and consumption pattern of alcohol in a particular state which can only be studied in depth provided a sufficient national data eco-system.

## Data Availability

Data can be shared publicly on request. Please email the corresponding author for the data.

## Footnotes

3 In Appendix-1, we develop an econometric model of determinants of alcohol consumption where the determinants are household characteristics, *X*_*h*_.

4 See: About the SAFER initiative to reduce alcohol related harm (who.int)

